# SARS-CoV-2 pandemic dynamics and infection tracing in Denmark

**DOI:** 10.1101/2021.03.15.21253582

**Authors:** Morten W.N. Jørgensen, Niels Høiby, Hans J. Ziock, Steen Rasmussen

## Abstract

We model and simulate the COVID-19 infection and healthcare dynamics in Denmark from the onset till March 5, 2021. The simulation is matched and calibrated to hospital and death data as well as antibody population measurement. In this work we focus on comparing the time evolution of the estimated infection level with the daily identified infected individuals based on the national testing and contact tracing program. We find that the national testing program on average identifies 1/3 of the infected individuals July 1, 2020 - March 5, 2021. Our investigations indicate the current program does not have a proper balance between random probing, focused contact tracing, and testing prioritization. Too much of the program operates as a semi-random daily sampling of part of the population. We propose a policy with a focus on local infection tracing and interventions.

## Introduction

We model and simulate the COVID-19 infection and healthcare dynamics in Denmark from the onset till March 5, 2021. The simulation is matched and calibrated to hospital and death data. For details see Ref. [1]. In this work we compare the time evolution of the estimated infection level with the daily identified infected individuals based on the national testing and contact tracing program.

### The model system

The model is built from a basic SEIRS (Susceptible – Exposed - Infectious - Recovered - Susceptible) style model and assumes people can be in one of the three infectious states: incubation *I*_*i*_ (no symptoms), asymptomatic *I*_*a*_ (no or only weak symptoms), and symptomatic *I*_*s*_, see Fig. 1.

**Figure 1.**
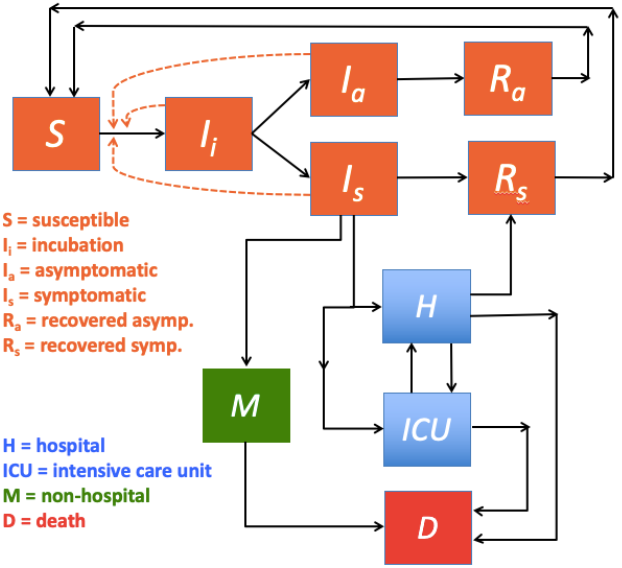
Flow diagram of the SEIRS based model system. As only ∼3% are vaccinated by March 5, 2021 they are not explicitly modeled.

Upon infection, a fraction of the symptomatic population *I*_*s*_ can become seriously ill and are either transferred to a hospital *H* or an *ICU*. From the regular hospital unit *H* patients can recover, move to an *ICU*, or die *D*. Some individuals also die outside of the hospital system, mainly in homecare facilities. We have two categories of recovered as the duration of immune memory depends on the severity of the infection. In Fig. 1, a flow diagram of the infection and healthcare dynamics is shown. See [1] for details on the differential equation system and parameter values that govern the pandemic dynamics.

## Method

The scale of the infection is mainly determined by infection parameters *ß*_*x*_, *x* = *i, a, s* for the three different infection groups *I*_*i*_, *I*_*a*_, and *I*_*s*_, where *I*_*s*_ is most infectious, *I*_*a*_ second, and *I*_*i*_ least. Closing and the reopening of the country is modeled by modulating the *ß*’s, i.e., by initially decreasing the *ß*’s starting at the closedown dates, followed by increasing *ß*_*i*_ and *ß*_*a*_ after the dates where the national reopening policies change. We assume almost all *I*_*s*_ are isolated either at home or at hospitals, so the pandemic is mainly driven by the *I*_*i*_ and *I*_*a*_ populations. The historical succession of closings and reopenings of Denmark is shown in Fig. 2, where we modulate the impact on the *ß*’s to match the national hospital and *ICU* occupations, as well as death data. Once the *ß* values are fixed, we can calculate *ℜ*_*0*,_ the reproduction number of the pandemic. All estimated *ß* and *ℜ*_*0*_ values are shown in Fig. 2.

**Figure 2.**
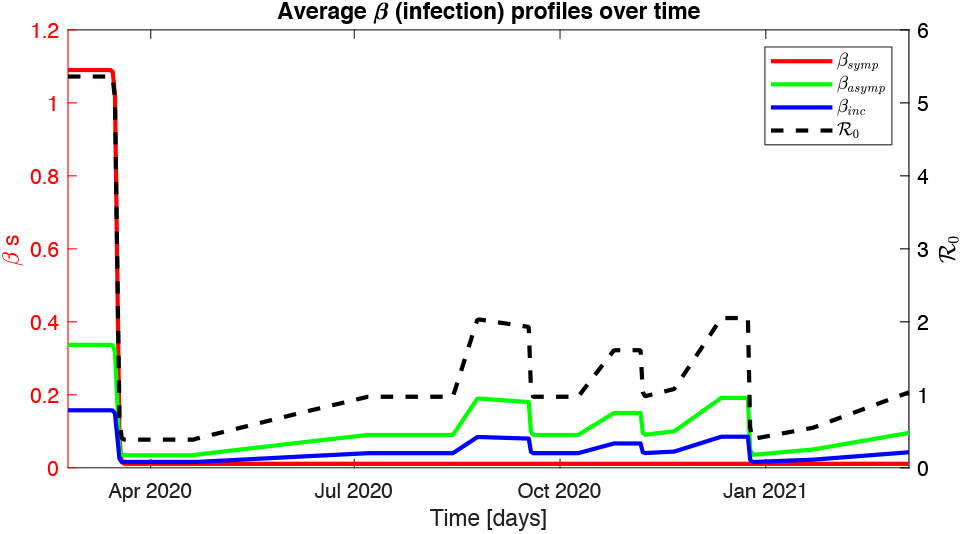
*ß* and *ℜ*_*0*_ profiles. Initial free pandemic is followed by an initial effective lockdown of Mar. 11, 2020. Slow controlled reopenings in late spring and early summer are followed by opening of bars and nightlife by mid-August. A second boost occurs mid fall and infections continue to increase due to winter and Christmas followed by a 2^nd^ full lockdown by Dec. 25. The steady increase thereafter is due to the increasing B117 mutant and slow reopening late Feb. 2021.

## Results

We compare the reported and simulated hospital and death data in Fig. 3. It should be emphasized that it is non-trivial to optimize the model parameters so simulations generate matching values. Parameters are in part identified from clinical studies^[2]^, duration of hospitalizations^[2]^, and antibody measurements^[3]^. For more details see Ref. [1]. For our infection tracing analysis, it is important to note the size of our estimated symptomatic versus asymptomatic ratio value. Combining model and national antibody measurement data^[3]^ yield about 1 in 5 infected with clear symptoms (fever). Thus, by far, most infected are asymptomatic as also supported by clinical studies^[8] [9] [10]^.

**Figure 3.**
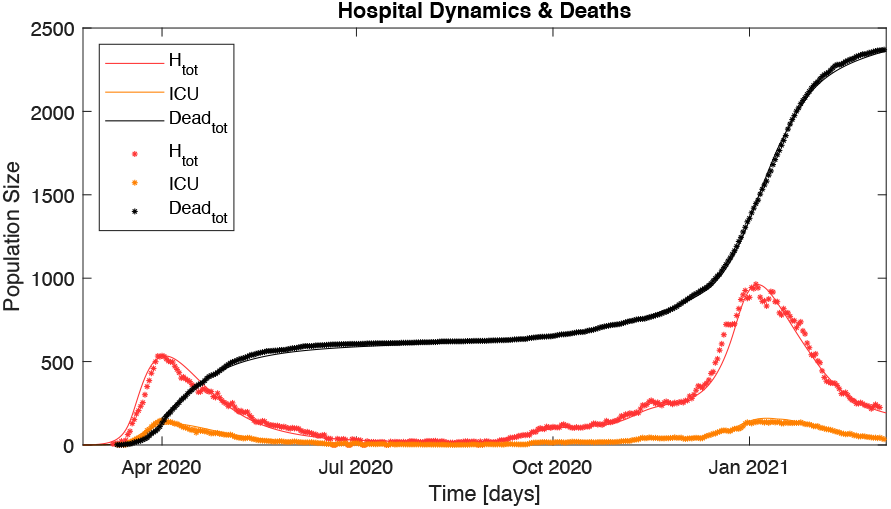
Comparison of reported (dots) and simulated (solid lines) daily hospital (*H* + *ICU*) and *ICU* occupation values, as well as total death data both inside and outside the health care system.

Fig. 4 shows the national data for the daily infection tracing^[7]^ together with the simulated data for the infected population *I*_*tot*_ = *I*_*i*_ + *I*_*a*_ + *I*_*s*_ from July 1, 2020 onwards. Note a more than doubling in infection tracing from July to August. Also note the significant infection tracing peak (Dec. 16, 2020) just before Christmas, where many people were tested so that they could enjoy holidays with their families, while the infection curve continues to increase until Dec. 25, 2020 when a strict lockdown was reimposed. A steep increase in the infection tracing is also seen in late Oct., where an early discovered, mink farm driven, major local outbreak started in the Northern Jylland region. Almost everybody in the region was tested. The region was under lockdown and the outbreak was contained. In other regions there was subsequently also a rise of covid-19 cases perhaps sparked by the one-week school holiday in Oct. coinciding with colder weather and more indoor activities.

**Figure 4.**
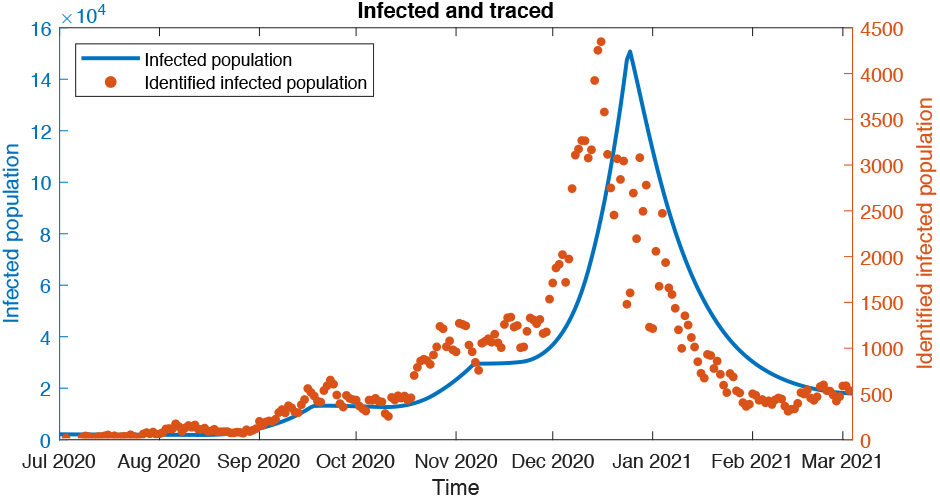
Time evolution of simulated *I*_*tot*_ (solid blue) and daily reported infection tracing^[7]^ (red dots) based on the national testing and contact tracing program. Note different scales. See text for details.

The daily efficiency of the national testing and contact tracing program (*DT*) may be estimated by comparing the actual daily identified infected *T*_*d*_(*t*) with our simulation based estimated daily number of infected given by *I*_*i*_ and *I*_*s*_. We assume an average incubation time 1/*γ* = 5 days and the average duration time as symptomatic and asymptomatic to be 1/*γ*_*s*_ = 1/*γ*_*a*_ = 10 days. Further, the incubated on average start to become infectious the last 1.5 days of their incubation time.

If the positive diagnosis occurs on the day they become infected, we can define *DT*^*max*^(t) = *T*_*d*_(*t*) / (*I*_*i*_(*t*)*γ*_*i*_). If the identification occurs as late as possible in their disease process, we can define *DT*^*min*^(*t*) = [*f*_1_*I*_*s*_(*t*)*γ*_*s*_ + *f*_2_(*T*_*d*_(*t*) – *I*_*s*_(*t*)*γ*_*s*_)] / (*I*_*i*_ (*t*)*γ*_*i*_,). Here *f*_1_ = 10.0/11.5 defines the fraction of time an infected who eventually becomes symptomatic is prevented from infecting, while *f*_2_ = 1.0 /11.5 defines the fraction of time an infected who eventually becomes asymptomatic is prevented from infecting. The above assumes that all symptomatic infected are identified once they become symptomatic.

As *DT*^*max*^(*t*) and *DT*^*min*^(*t*) define upper and lower limits for the infection detection rates, and we don’t have information on when within their time as infected they are identified, a best (maximum entropy) estimate for the actual daily detection rate is *DT*^*avg*^(*t*) = [*DT*^*max*^(*t*)+ *DT*^*min*^(*t*)]/2, see Fig 5.

**Figure 5.**
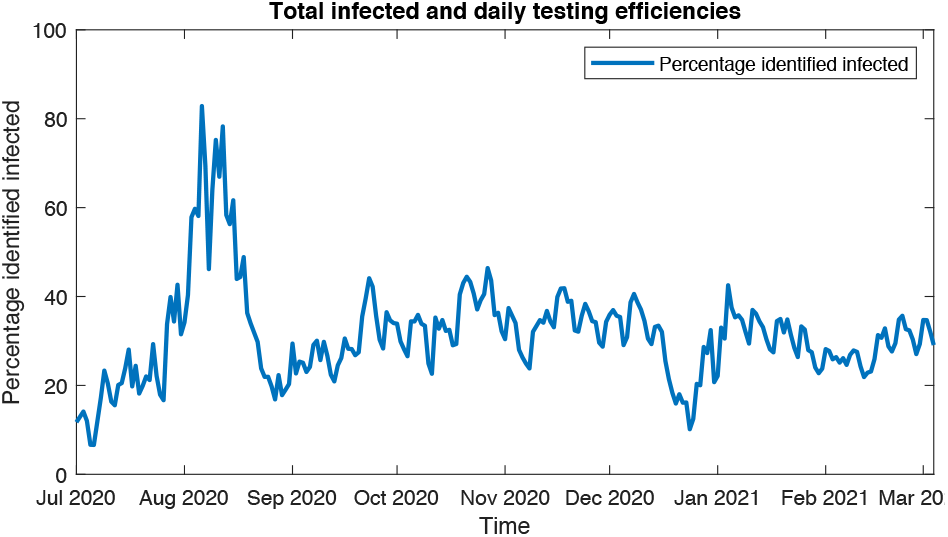
Efficiency of daily infection tracing in Denmark Jul. 1, 2020 – Mar. 5, 2021 expressed as *DT*^*avg*^(*t*). See text for details.

## Discussion and Conclusions

We have expanded and calibrated the COVID-19 simulation from Ref. [1] through Mar. 5, 2021 mainly based on hospital and death data; recall discussions around Figs. 1-3. Simulation estimates for the size of the infected population each day can now be compared with the daily reported infection tracing. From these two numbers, together with details about the nature of the pandemic, we can estimate the efficiency of the daily infection tracing, which is discussed around Figs. 4 and 5. It should be noted that our estimates are by nature associated with uncertainties, as there is at present no directly way to measure the actual scale of the infected population size.

We estimate an average infection tracing efficiency of around 1/3 from Sept. 2020 through Feb. 2021. The large efficiency-increase in Aug. 2020 stems from a highly successful local infection tracing effort in the Aarhus region of Denmark, where a local outbreak was stopped before it could spread to other regions. It should be noted that the large scale of the fluctuations is due to the very low macroscopic national infection level at that time.

Denmark has an ambitious testing program, currently a daily capacity of about 5% of the population. However, our investigations indicate the current program does not have a proper balance between random probing, focused contact tracing, and testing prioritization. Too much of the program operates as a semi-random daily sampling of part of the population. Two examples of documented successful focused infection tracing intervention occurred in the Aarhus and Northern Jylland regions, which could act as models. Note the Northern Jylland success is not directly visible in Fig. 5 because of the much higher national infection level at that time.

## Data Availability

All used data are publicly avilable and pointers to data are clearly stated in the references of the paper.

https://www.ssi.dk

https://www.stps.dk

https://www.dr.dk

## Acknowledgement

We are grateful to Michael S. Petersen for constructive discussions and suggestions for this work, and Emilie M. Andersen and Søren V. Iversen for helping with the initial infection tracing simulations as part of their SDU course work on mathematical modeling and simulation.

## Notes

### Competing Interest Statement

The authors have declared no competing interest.

### Funding Statement

No external funding was received (no funding from third parties). Funding was provided by independent university salaries.

### Author Declarations

We have followed all appropriate reseatch reporting guidelines

